# Intra-host site-specific polymorphisms of SARS-CoV-2 is consistent across multiple samples and methodologies

**DOI:** 10.1101/2020.04.24.20078691

**Authors:** Rebecca Rose, David J. Nolan, Samual Moot, Amy Feehan, Sissy Cross, Julia Garcia-Diaz, Susanna L. Lamers

## Abstract

Despite the potential relevance to clinical outcome, intra-host dynamics of SARS-CoV-2 are unclear. Here, we quantify and characterize intra-host variation in SARS-CoV-2 raw sequence data uploaded to SRA as of 14 April 2020, and compare results between two sequencing methods (amplicon and RNA-Seq). Raw fastq files were quality filtered and trimmed using Trimmomatic, mapped to the WuhanHu1 reference genome using Bowtie2, and variants called with bcftools *mpileup*. To ensure sufficient coverage, we only included samples with 10X coverage for >90% of the genome (n=406 samples), and only variants with a depth >=10. Derived (i.e. non-reference) alleles were found at 408 sites. The number of polymorphic sites (i.e. sites with multiple alleles) within samples ranged from 0-13, with 72% of samples (295/406) having at least one polymorphic site. Correlation between number of polymorphic sites and coverage was very low for both sequencing methods (R^2^ < 0.1, p < 0.05). Polymorphisms were observed >1 sample at 66 sites (range: 2-38 samples). The minor allele frequency (MAF) at each shared polymorphic site was 0.03% - 48.5%. 33/66 sites occurred in ORF1a1b, and 37/66 changes were non-synonymous. At 10/66 sites, derived alleles were found in samples sequenced using both methods. Polymorphic amplicon samples were found at 10/10 positions, while polymorphic RNA-Seq samples were found at 7/10 positions. In conclusion, our results suggest that intra-host variation is prevalent among clinical samples. While mutations resulting from amplification and/or sequencing errors cannot be excluded, the observation of shared polymorphic sites with high MAF across multiple samples and sequencing methods is consistent with true underlying variation. Further investigation into intra-host evolutionary dynamics, particularly with longitudinal sampling, is critical for broader understanding of disease progression.

## MAIN TEXT

Researchers worldwide have engaged in a remarkable effort to generate and share genomic sequences of SARS-CoV-2, in particular to the GISAID database (gisaid.org), resulting in nearly 10,000 temporal and geo-located whole genomes available in mid-April 2020 [1]. These data, and sophisticated methods, have enabled a robust estimation of a relatively slow population evolutionary rate (∼0.8 × 10-3 substitutions per site per year, [2]). As additional genomes are uploaded each day, viral dynamics on a population level can be refined in real-time.

On the other hand, a similarly thorough investigation of the intra-host evolutionary rate has yet to be published, resulting in part from the scarcity of patient longitudinal samples. In addition, analyzing published datasets of raw sequence data is much more time-consuming than using consensus genomes, and the lack of a common sequencing protocol and analytical pipeline renders comparisons among samples difficult. Finally, much less raw data have been made available: for example, as of 14 April 2020, <1,000 raw sequence files were available from the Short Read Archive (SRA) at the National Center for Biotechnology Information (NCBI).

Despite these challenges, evidence from numerous angles suggests a potentially important role of intra-host variation. For example, a few studies have demonstrated the presence of intra-host variation of SARS on a population level, and more intriguingly, variation occurring at the same position in multiple individuals [3, 4]. These observations suggest that either that patients are becoming infected with a similarly diverse viral population, or that the virus is following a similar evolutionary trajectory in multiple patients, both of which may have clinical implications. Furthermore, the evolution of shared variants has a well-established impact on viral replication and tropism in both human viruses (e.g. HIV-1) and other coronaviruses (e.g. feline).

Our goal in this study was to quantify and characterize intra-host variation in all SARS-CoV-2 raw sequence data available in SRA as of 14 April 2020, and compare results between two sequencing methods (amplicon and RNA-Seq).. A search for “SARS-CoV-2” resulted in a total of 1173 samples. We downloaded all samples that were classified as “Wuhan seafood market pneumonia virus” or “Severe acute respiratory syndrome coronavirus 2”; host = *Homo sapiens;* derived from a primary isolate; sequenced using an Illumina platform, and downloaded successfully, resulting in a total of 441 samples (**Supplemental File 1**).

Raw fastq files were quality filtered and trimmed using Trimmomatic and mapped to the WuhanHu1 reference genome (NC_045512.2) using Bowtie2. Since the reference sequence was one of the earliest samples from geographic origin of the current pandemic, this was considered the ancestral sequence. Bam files were sorted and indexed with samtools, and variants called with bcftools *mpileup*. Only variants with a depth >=10 were retained. Individual vcf files were merged, and allele frequencies were extracted using the *vcfR* package in R. The complete commands are available in **Supplemental File 2**.

While all raw data was generated using the Illumina short read technology, numerous experimental variables differed among studies, (e.g. assay type, library construction kit, instrument, etc). We sorted the dataset into two general groups based on their sequencing approach: those which sequenced just PCR amplicons of SARS-CoV-2 vs. those that used a general RNA-Seq metagenomic method which typically sequences all the RNA isolated from a sample unless specific RNA classes are excluded i.e. rRNA depletion, or enriched i.e. mRNA selection. Since an accurate estimation of intra-host variation requires sufficient genomic coverage, we first investigated sample quality by visualizing mapping metrics for all samples.

In general, coverage was higher for the amplicon data compared with the RNA-Seq data (**Supplemental Figure 1**). The interquartile range of mean coverage was 239 – 246 for the amplicon data, and 9-231 for the RNA-Seq data. The percent of the genome covered at 1x was high for both (amplicon IQR: 0.997 – 0.998; RNA-Seq IQR: 0.916 – 0.999). However, while the percent of the genome covered at 10x and 25x remained high with the amplicon data (10x IQR: 0.987 – 0.997; 25x IQR: 0.980 – 0.997), the range was much broader for the RNA-Seq data (10x IQR: 0.240 – 0.997; 25x IQR: 0.024 – 0.993). This suggests that, particularly for studies investigating intra-host variation, amplicon-based sequencing may be preferable.

**Figure 1.**
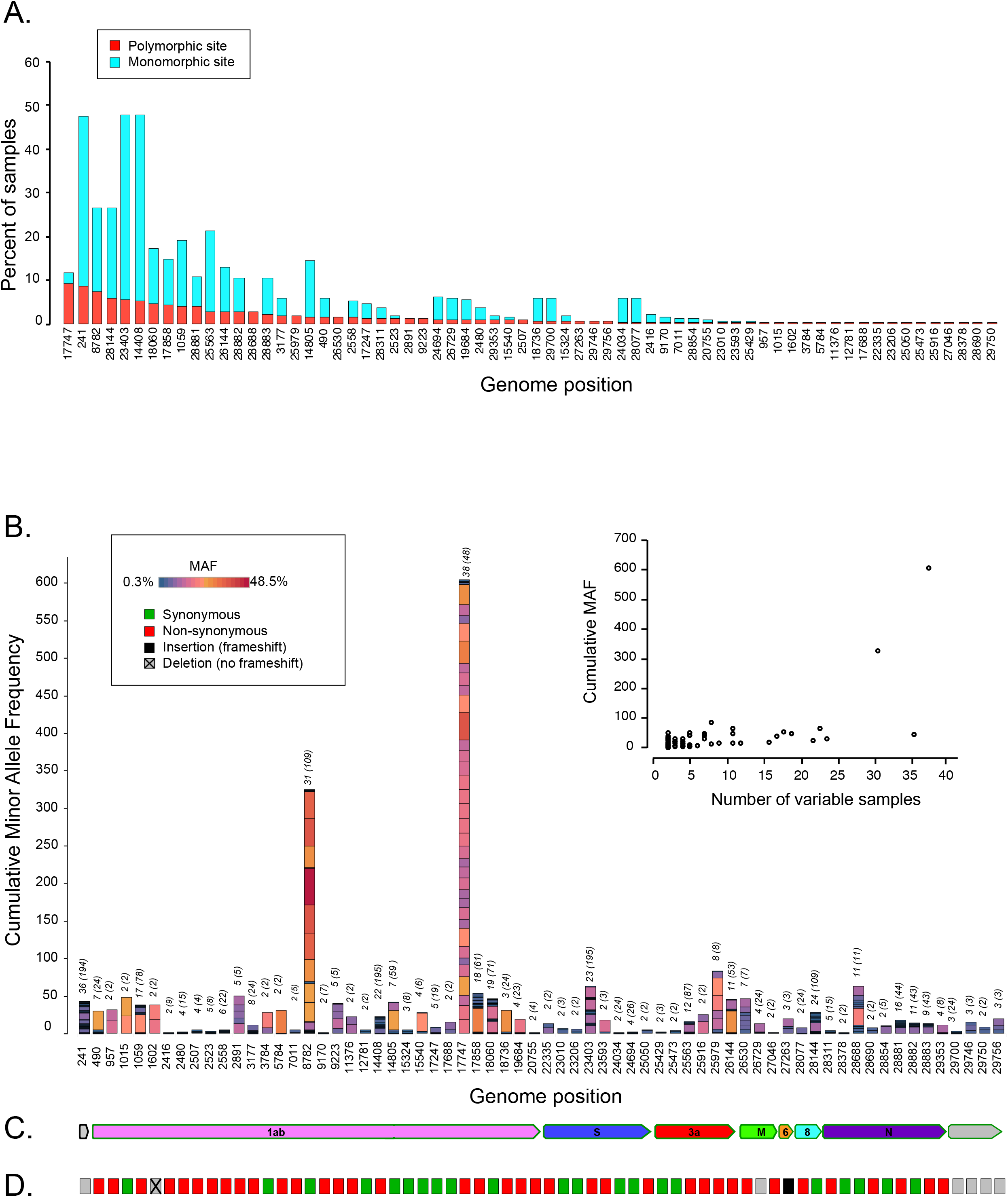
Characteristics of 66 shared polymorphic positions of the SARS-CoV-2 genome. **(A)** Percent of samples (y-axis) with the derived variant out of the total number of samples (n=406) at each position in the genome (x-axis); positions are sorted by MAF. Red bars = polymorphic samples; blue bars = monomorphic samples. (**B**) Cumulative minor allele frequency (MAF; y-axis) at each genome position (x-axis); positions are sorted by order in genome. The color and size of each block making up the columns represents the MAF of one sample according to the legend. The number of polymorphic samples for each position is indicated by the number above the column; the number in parentheses indicates the total number of samples with the derived allele at each position. (**C**) Colored arrows represent SARS-CoV-2 genes and are placed with respect to (**B)** to indicate the genes in which each polymorphic position occurs. (**D**) Each colored box corresponds to a genome position in **(B**). Boxes are colored according to the legend and indicate the predicted impact of the derived allele on the amino acid.

To ensure sufficient coverage, we excluded samples with 10X coverage for <90% of the genome (11/41 RNA-Seq samples and 21/400 amplicon samples), resulting in a retained total of 406 samples (**Supplemental File 3**). Among these 406 samples, derived (i.e. non-reference) alleles were found at 408 positions across the genome (**Supplemental File 4**). The number of sites with the derived allele per sample ranged from 0-15. The number of polymorphic sites (i.e. sites with multiple alleles) within samples ranged from 0-13, with 72% of samples (295/406) having at least one variable site. Correlation between number of polymorphic sites and mean/10x coverage was very low for both sequencing methods (R^2^ < 0.1, p < 0.05 for all cases, **Supplemental Figure 2**).

We then assessed how polymorphic sites were distributed among samples. 294/406 positions were polymorphic in at least one sample, while 66 positions were polymorphic in >1 sample. For each site, the percentage of polymorphic samples ranged from 9.3% (38/408) for position 17747 to 0.05% (4/408) for 27 different positions (**Figure 1A**). Across all 66 positions, the weighted proportion of polymorphic samples compared to all samples with the derived allele was 0.43. Interestingly, position 17747 had both the highest number of polymorphic samples (38/408) as well as the highest proportion of polymorphic samples (0.80).

Next we looked the minor allele frequency (MAF) in each sample for all of the 66 shared polymorphic sites (**Figure 1B**). MAF per site/per sample ranged from 0.03% to 48.5%. The combined MAF for all polymorphic samples per site ranged from 0.08% to 604%. There was a moderate correlation between combined MAF and the number of polymorphic samples (R^2^ = 0.43; **Figure 1B**). However, two sites in particular were notable for high MAFs among multiple samples: 8782 and 17747. When these sites were removed, the correlation was reduced (R^2^ = 0.23).

Of the 66 shared polymorphic positions, 50% occurred in ORF1a1b, including the two sites with the highest MAFs (8782 and 17747). The remaining positions broke down as follows: ORF S: n=8; ORF 3a: n=6; ORF M: n=3; ORF6: n=1; ORF8: n=2; ORF N: n=9 (**Figure 1B; Supplemental File 5)**. Five positions occurred in either the 5’ or 3’ UTRs. Of the 61 polymorphic positions occurring in translated region, 59 were single nucleotide polymorphisms (SNPs), of which 37 were non-synonymous (including position 17747), and 22 were synonymous (including positions 8782). Two derived alleles at polymorphic positions were deletions, one of which retained the reading frame, while the other did not.

Finally, to determine the potential effect of sequencing method on intra-sample variation, we plotted the MAF and the number of samples with a derived variant for each of the 66 shared polymorphic sites (**Supplemental Figure 3**). The majority of polymorphic sites included samples which were sequenced using the amplicon methodology, reflecting the much more common use of amplicon sequencing in this dataset. At 10 sites, derived alleles were found in samples sequenced using both methods (241, 8782, 14408, 14805, 17747, 17858, 18060, 23403, 26144, and 28144). Polymorphic amplicon samples were found at all 10 positions, while RNA-Seq processed samples were found at 7/10 positions. At the two positions with the highest MAFs (8782 and 17757), polymorphic samples were found in both methods, although clearly higher among the amplicon processed samples.

In conclusion, our results suggest that intra-host variation is prevalent among clinical samples, most of which is masked when constructing consensus genomes. While this variation could be due to amplification and/or sequencing errors, evidence supporting real virus variation includes nonrandom distribution of polymorphic sites, shared polymorphic sites among up to 38 samples; MAF up to 50%; and detection of polymorphisms at the same site using two very different sequencing methods. The high number of mutations that change the amino acid suggest a potential functional relevance. Further investigation into intra-host evolutionary dynamics, particularly with longitudinal sampling, is critical for broader understanding of disease progression.

## Data Availability

All raw data is available in the SRA database. Results and scripts are provided as supplemental material.

## Acknowledgments

RR, DN, SC, SM, and SLL are employed by Bioinfoexperts, LLC. The authors thank all researchers for making raw data publicly available.

## Financial declaration

SLL is the recipient of National Science Foundation SBIR funding (#1830867).

## Figure Legends

**Supplemental Figure 1. Boxplots of mean coverage, percent genome coverage at 1X, 10x and 25x for 441 samples by sequencing method**. Dots represent samples; interquartile range indicated by grey box and noted above each figure. Whiskers extend to 1.5x the IQR.

**Supplemental Figure 2**. Scatterplot of number of polymorphic sites vs. mean coverage and 10x coverage. Dots represent 406 samples, colored by sequencing method (red = RNA-Seq; blue = amplicon). Regression lines and R^2^ value are shown for each comparison.

**Supplemental Figure 3. Minor allele frequencies for 66 shared polymorphic positions. Each dot indicates the MAF (y-axis) for each of 66 positions (x-axis)**. Dots are colored to indicate polymorphic (red) or monomorphic (i.e. MAF = 0; blue) sample. Samples are separated by experimental method. Asterisks are placed above the 10 positions with the derived allele found in samples processed with both methods; a single asterisk indicates all RNA-Seq processed samples were monomorphic; double asterisk indicates >=1 RNA-Seq processed samples were polymorphic.

